# The Cardiovascular Manifestations and Management Recommendations for Ogden Syndrome

**DOI:** 10.1101/2025.02.11.25321331

**Authors:** Rikhil Makwana, Rahi Patel, Rosemary O’Neill, Elaine Marchi, Gholson J. Lyon

**Affiliations:** Department of Human Genetics, New York State Institute for Basic Research in Developmental Disabilities, Staten Island, New York, United States of America; George A. Jervis Clinic, New York State Institute for Basic Research in Developmental Disabilities, Staten Island, New York, United States of America; Biology PhD Program, The Graduate Center, The City University of New York, New York, United States of America

## Abstract

The NatA complex is composed of the NAA10, NAA15, and HYPK subunits. It is primarily responsible for N-terminal acetylation, a critical post-translational modification in eukaryotes. Pathogenic variants within *NAA10* cause Ogden Syndrome (OS), which is characterized by varying degrees of intellectual disability, hypotonia, developmental delay, and cardiac abnormalities. Although the cardiac manifestations of the disease have been described extensively in case reports, there has not been a study focusing on the cardiac manifestations and their recommended clinical cardiac management. In this study, we describe the cardiac manifestations of OS in a cohort of 85 probands. We found increased incidence of structural and electrophysiologic abnormalities, with particularly high prevalence of QT interval prolongation. Sub-analysis showed that male probands and those with variants within the NAA15-binding domain had more severe phenotypes than females or those with variants outside of the NAA15-binding domain. Our results suggest that an OS diagnosis should be accompanied by full cardiac workup with emphasis on echocardiogram for structural defects and EKG/Holter monitoring for electrophysiologic abnormalities. Additionally, we strongly recommend that the use of QT-prolonging drugs be followed up with routine electrophysiological monitoring or consultation with a pediatric cardiologist. We hope this study guides clinicians and caregivers treating patients with OS and moves the field toward a standardized diagnostic workup for patients with this condition.

## Introduction

N-α-terminal acetylation is a highly conserved form of co-translational protein modification whereby an acyl group is transferred from Acetyl-CoA to the α-amino group at the N-terminus of a protein in order to modify the half-life, structure, or localization of the final product ^1–7^. The most prevalent of N-α-terminal acetyltransferases is the NatA complex which targets around 40% of the human proteome^1,8^. The NatA complex is composed of three distinct sub-units: NAA10, NAA15, and HYPK^9,10^. NAA10 functions as the catalytic domain, NAA15 as an auxiliary domain, and HYPK as a regulatory subunit^11–13^.

*NAA10* is considered an essential human gene as knockouts of it are incompatible with life^14^. *NAA10*’s transcript is 235 amino acids (AA) in length and includes a NAA15 interaction domain and a Gcn5-related N-acetyltransferase (GNAT) domain^10^. Variants in *NAA10* have been implicated in various cancers including non-small cell lung cancer^15,16^, renal cell carcinoma^17,18^, squamous cell carcinoma of the esophagus^19,20^, colon cancer^21,22^, and more^23–28^.

*NAA10* variants also lead to a distinct clinical phenotype characterized primarily by intellectual disability, hypotonia, variable developmental delay, dysphagia, difficulty feeding, and variable ophthalmic manifestations known as NAA10-related neurodevelopmental syndrome or Ogden Syndrome (OS)^29–32^. OS is an X-linked ultra-rare genetic syndrome. The first two families were sequenced and reported in 2011^33^. In addition to the effects on the nervous, gastrointestinal, and ophthalmic systems, *NAA10* variants lead to various cardiac manifestations that have been previously described in case reports, such as cardiomyopathy^34–38^ and prolonged QT intervals^35,39,40^. Variants in the autosomal gene *NAA15* can also present with variable developmental delay and cardiac manifestations, these individuals tend to have much less severe manifestations than those with OS^29,31,32,41–45^.

Specific cardiomyocyte models of OS disease have been developed through the creation of patient derived iPSC cell lines of the p.S37P (severe) and p.Y43S (milder) variants. These studies suggested there may exist specific *NAA10*-related arrythmias secondary to increased calcium conductance through L-type voltage gate calcium channels^46^. Cell lines from a patient with the p.R4S were also developed and showed similarly increased calcium conductance, as well as sodium and potassium conductance dysfunction and cytoarchitectural and contractile abnormalities, as reported in a preprint^42^.

There have been several case reports and manuscripts describing the genotype-cardiac phenotype relationship of singular or familial cases of OS. However, there has not yet been an overarching study focusing on a large cohort of individuals with OS and their cardiovascular manifestations of disease. Through a more thorough exploration of the cardiac manifestations of OS, we hope to provide direct guidance to clinicians and caregivers treating a child with OS. Additionally, we hope to expand the current understanding of *NAA10* variant clinical manifestations.

## Methods

### Participant Data

The subject population is composed of a group of probands diagnosed with OS who have either previously worked with the principal investigator G.J.L. or who have been referred to him for his expertise on the condition. Probands and their caregivers signed Institutional Review Board-approved consent forms and HIPAA forms to allow for their information to be used in the study, with approval of protocol #7659 for the Jervis Clinic by the New York State Psychiatric Institute— Columbia University Department of Psychiatry Institutional Review Board, followed by recent transfer to and re-approval by the Nathan Kline Institute Institutional Review Board (in 2024). The scope of the project was explained in lay language, and there was no financial compensation offered for participation.

Thorough medical histories were collected on all probands, including genetic testing results and any cardiac or pulmonary-specific histories or testing (EKG, echocardiogram, cardiac MRI, Holter monitors, etc.). Medical records were either directly received from probands/caregivers or collected through direct requests from their treating hospitals, with patient permission. Clinical exome sequencing was used to confirm the pathogenicity of each variant. All data was organized based on a unique identifier system, “OS_XXX”, that functions as an internal registry of individuals with Ogden Syndrome. This registry and its key are known only to the research team. All proband medical data related to cardiac function was aggregated into an Excel spreadsheet and reviewed for accuracy by two independent members of the research team. Narrative histories were annotated and probands were characterized as having the presence or absence of cardiac pathology. For a patient to be classified as having a cardiac pathology for the purposes of this study, an official diagnosis needed to be present on their associated medical records. Automated diagnosis, such as those that accompany EKG readings, were not sufficient to classify the presence or absence of pathology unless this interpretation was confirmed by two independent members of the research team or documented by the proband’s cardiologist or provider. Additionally, structural abnormalities were characterized discretely as either present or absent without grading.

Pathologies were grouped grossly into 6 distinct categories: structural, valvular, myocardial, pericardial, vascular, and cardiac electrophysiologic. Structural abnormalities include patent foramen ovale (PFO), atrial septal defect (ASD), ventricular septal defect (VSD), patent ductus arteriosus (PDA), bicuspid aortic valve, aortic root dilatation, ascending aorta dilation, over-riding aorta, and tetralogy of Fallot. Valvular abnormalities include tricuspid regurgitation (TR), mitral regurgitation (MR), pulmonic regurgitation, aortic stenosis, and mitral stenosis. Myocardial pathologies include hypertrophic cardiomyopathy (HOCM), non-descript ventricular hypertrophy, and cardiomegaly. Pericardial abnormalities include pericardial effusion, pericarditis, and apical cyst. Vascular abnormalities include persistent left superior vena cava (PLSVC), pulmonary hypertension, pulmonary vessel stenosis, arterial stenosis, and coronary fistula. Electrophysiologic abnormalities include bradycardia, shortened PR interval, prolonged QT interval, supraventricular tachyarrhythmias (SVT), AV nodal re-entrant tachycardia (AVNRT), atrial fibrillation, ventricular fibrillation, and ventricular tachycardia. Additional cardiac pathology observed and included in total pathology counts but not in the broader categories include syncopal episodes, dextrocardia, cardiac arrest, myocardial infarction, and heart failure.

### Analysis

All deidentified data was compiled into an Excel spreadsheet stored in a HIPAA compliant drive. Descriptive statistics, including demographic information, discrete counts of proband cardiac pathology, average and standard deviation (SD) of EKG parameters, and the types and number of cardiac tests received were compiled for each proband. EKG parameters analyzed included rate, rhythm, and length of the PR, QRS, and QT intervals in milliseconds (ms). QTc for each EKG was calculated via the Bazett formula^48^. To ensure that there was no overcounting, the mean rate, PR, QRS, QT, and QTc values were used for probands with greater than one EKG reading. Additionally, due to incomplete records, not all electrophysiological data included all components of the EKG such that certain probands were included in calculations for only the rate, only PR interval and rate, etc. Current ages were calculated by the probands’ date of birth subtracted from the date of the data freeze performed on November 22, 2024. For those probands who are deceased, their age at the time of death was calculated.

Analyses were repeated by proband sex and by the presence of a variant within the NAA15 interaction domain (AA 1-58). One-tailed unequal variance t-tests were calculated comparing the prevalence of each category of cardiac pathology and total number of pathologies. Two-tailed unequal variance t-tests were calculated comparing the rate and length of rate, PR, QRS, QT, and QTc values. Chi square statistics were calculated comparing each group’s presence or absence of ever having a recorded QTc interval greater than 440 ms in males or greater than 460 ms in females. Fisher’s exact test values were calculated comparing each group’s proportion of probands with at least one cardiac pathology and across all discrete counts of cardiac pathologies and for the sex distribution of probands with variants within the NAA15 interaction domain. All alpha values were set to 0.05. The null hypothesis was that there exists no differences between groups. Our proposed alternate hypothesis is that male probands and probands with variants within the NAA15 interaction domain have more cardiac pathologies and more aberrant EKG parameters.

Sub-analysis was performed by further splitting probands into three groups—male, female Arg83Cys, and female non-Arg83Cys. The three groups were compared with two-sample unequal variance t-tests to isolate the effect of the greater number of probands with Arg83Cys variants in this cohort. A Bonferroni correction was made to account for multiple hypothesis testing with a modified α of 0.02.

Lastly, correlation analysis was performed to uncover correlations between the amino acid residue position and number of cardiac pathologies, EKG rate, PR, QRS, QT, and QTc values, and QTc length with the age of EKG acquisition. For probands with greater than three EKGs over time (OS_198, OS_106, OS_107, OS_108, OS_109, OS_127, OS_138, OS_137, OS_178, OS_182, and OS_152), the QTc analysis was repeated. P-values were calculated for each correlation coefficient and α was set at 0.05. A Bonferroni correction was applied to the individual EKG analysis to account for the multiple hypothesis testing with a modified alpha of 0.005 (n = 11 probands with greater than 3 recorded EKGs). Simple linear regressions for analyses performed over time and over amino acid residue location were calculated to aid in data visualization.

## Results

There were 85 probands included in the study—65 female and 20 males. The average age of the cohort was 16.5 years old (SD = 17.2). The average age of the females was 16.1 (SD = 12.1) and males was 17.9 (SD = 27.8). Seven deceased individuals were also included in the study, 6 males and 1 female. Included in the cohort were three families. The first was a mother and her two sons—all with p.Tyr43Ser variants. The second was a mother and her two sons—all with p.Ile72Thre variants. The third family is a mother and son both with p.Arg116Gln variants. Probands were from 16 different countries, primarily the United States (n = 41) and the United Kingdom (n = 13). There were 25 unique variants across the probands with Arg83Cys being the most prevalent (n = 33/85). A complete breakdown of the variants can be seen summarized in **Table 1**. Further demographic breakdown by country is available in **Supplemental Table 1**.

**Table 1.** Variant Breakdown by Sex.

| Variant | Females | Males | Total |
| --- | --- | --- | --- |
| p.Asp10Gly | 1 | 1 | 2 |
| p.His16Pro | 1 | 0 | 1 |
| p.Cys17Gly | 0 | 1 | 1 |
| p.Tyr31Cys | 1 | 0 | 1 |
| p.His34Tyr | 0 | 1 | 1 |
| p.Ser37Pro | 0 | 4 | 4 |
| p.Tyr43Ser | 1 | 2 | 3 |
| p.Ile72Thr | 1 | 4 | 5 |
| p.Arg83Cys | 32 | 1 | 33 |
| p.Ala87Ser | 3 | 0 | 3 |
| p.Gln88Pro | 1 | 0 | 1 |
| p.Glu100Lys | 1 | 1 | 2 |
| p.Ala104Asp | 1 | 0 | 1 |
| p.Arg116Gln | 2 | 1 | 3 |
| p.Arg116Trp | 3 | 0 | 3 |
| p.His120Pro | 2 | 0 | 2 |
| p.Leu121Val | 1 | 0 | 1 |
| p.Ser123Pro | 1 | 0 | 1 |
| p.Leu126Arg | 1 | 0 | 1 |
| p.Phe128Leu | 7 | 0 | 7 |
| p.Phe128Ser | 1 | 0 | 1 |
| p.Met147Thr | 4 | 0 | 4 |
| p.Arg149Trp | 0 | 1 | 1 |
| p.Thr152Arg*6 | 0 | 2 | 2 |
| p.Glu181Ala*67 | 0 | 1 | 1 |
| **Total** | **65** | **20** | **85** |

The probands in this study underwent various types of medical testing to discern cardiac structure and function. The average age of our cohort at time of first documented cardiac screening test was 6.1 years old (SD = 9.2). From the cohort, 66 probands received a baseline echocardiogram (echo) or electrocardiogram (EKG) post-diagnosis. Abnormal EKG or echo readings were present in 46.48% (n = 33/71) of our cohort. Out of the probands with abnormal EKG or echo readings, 60.61% (n = 20/33) received a follow-up Holter monitor or Cardiac MRI. For probands who received advanced cardiac testing, 75.0% (n = 15/20) were still alive at the time of the data freeze.

There were 175 total cardiac anomalies accounted for from the cohort—93 identified in the females and 82 in the males. Structural abnormalities (n = 52, males (M) = 22, females (F) = 30) were the most prevalent, followed by electrophysiologic abnormalities (n =49, M = 21, F = 28). Pericardial pathologies were the least prevalent (n = 9, M = 3, F = 6). A complete quantitative count of the cardiac pathologies by group can be found pictorially in **Figure 1** and summarized in **Table 2**. The counts of each individual cardiac pathology can be seen in Supplemental Table 2.

**Figure 1.**
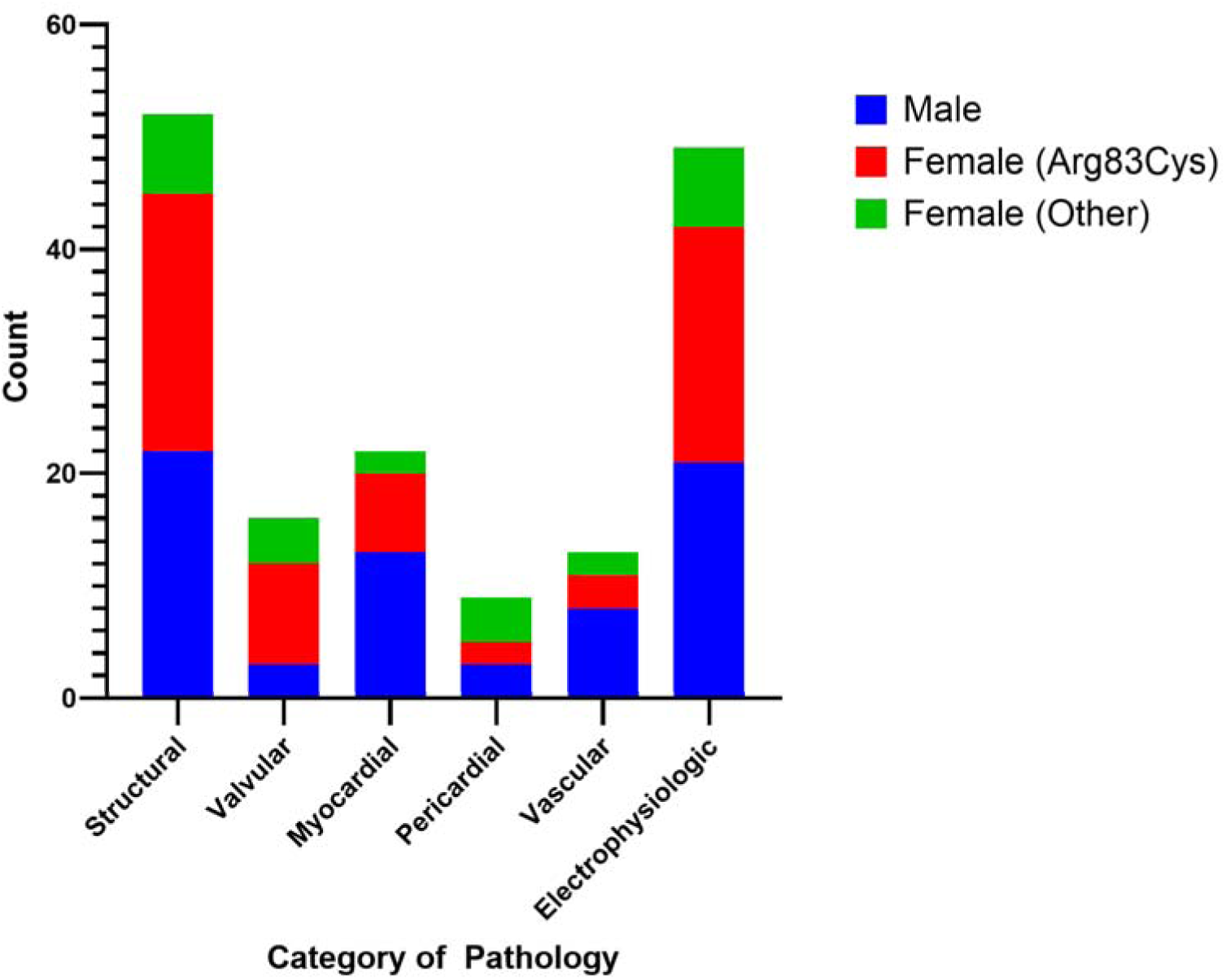
Discrete Number of Cardiac Pathology by Sex.

**Table 2.** Discrete Counts of Cardiac Pathology by Sex.

| Sex | Structural | Valvular | Myocardial | Pericardial | Vascular | Electrophysiologic | Total |
| --- | --- | --- | --- | --- | --- | --- | --- |
| Total Count | 52 | 19 | 22 | 9 | 13 | 49 | 178 |
| Male | 22 | 3 | 13 | 3 | 8 | 21 | 82 |
| Female | 30 | 13 | 9 | 6 | 5 | 28 | 93 |
| Female Arg83Cys<br>(R83C) | 23 | 9 | 7 | 2 | 3 | 21 | 67 |
| Female Other<br>(Other) | 7 | 4 | 2 | 4 | 2 | 7 | 26 |

The number of cardiac pathologies present in a single proband ranged from 0 (n = 30) to 12 (n = 1). Additional information on number of cardiac pathologies present can been seen in **Figure 2**. A breakdown of the number of probands with varying numbers of cardiac pathology broken down by sex and presence of a variant within the NAA15 interaction domain can been seen in **Table 3** and **Figure 3**. Males most often had four cardiac pathologies (35%, n=7/20), while females most often had zero cardiac pathologies (43.1%, n=28/65). Probands with variants within the NAA15-interaction domain of NAA10 also most often had four cardiac pathologies (35%, n = 7/20), while those with variants outside of it most often did not have any cardiac pathology (43.1%, n = 28/65). Furthermore, all probands with variants within the NAA15 interaction domain (n=13) had at least one cardiac pathology. There was a statistically significant greater proportion of probands with at least one cardiac pathology in the males compared to the females (p = 0.005) and in probands with variants within the NAA15 interaction domain (p = 0.002). There was also a statistically significant difference in the distribution of the gross number of cardiac pathologies when compared by sex (p = 0.0003) and by variant type (p = 0.0004).

**Figure 2.**
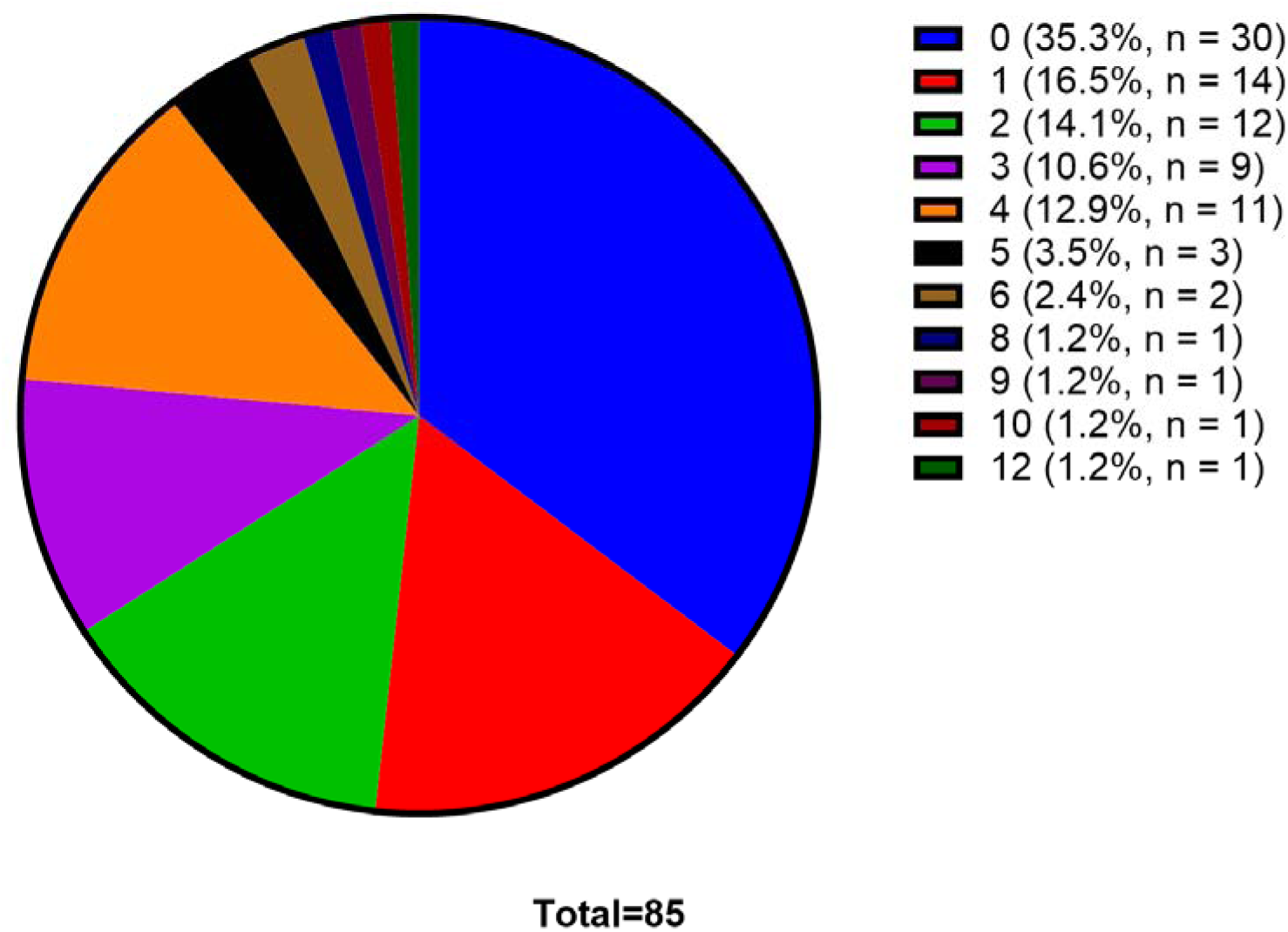
Breakdown of Probands by Number of Cardiac Pathology. Each section in the graph corresponds to the percentage of the total (n = 85) number of probands with that number of cardiac pathologies. The figure legend corresponds the number of cardiac pathologies to the percentage and discrete count of corresponding probands.

**Figure 3.**
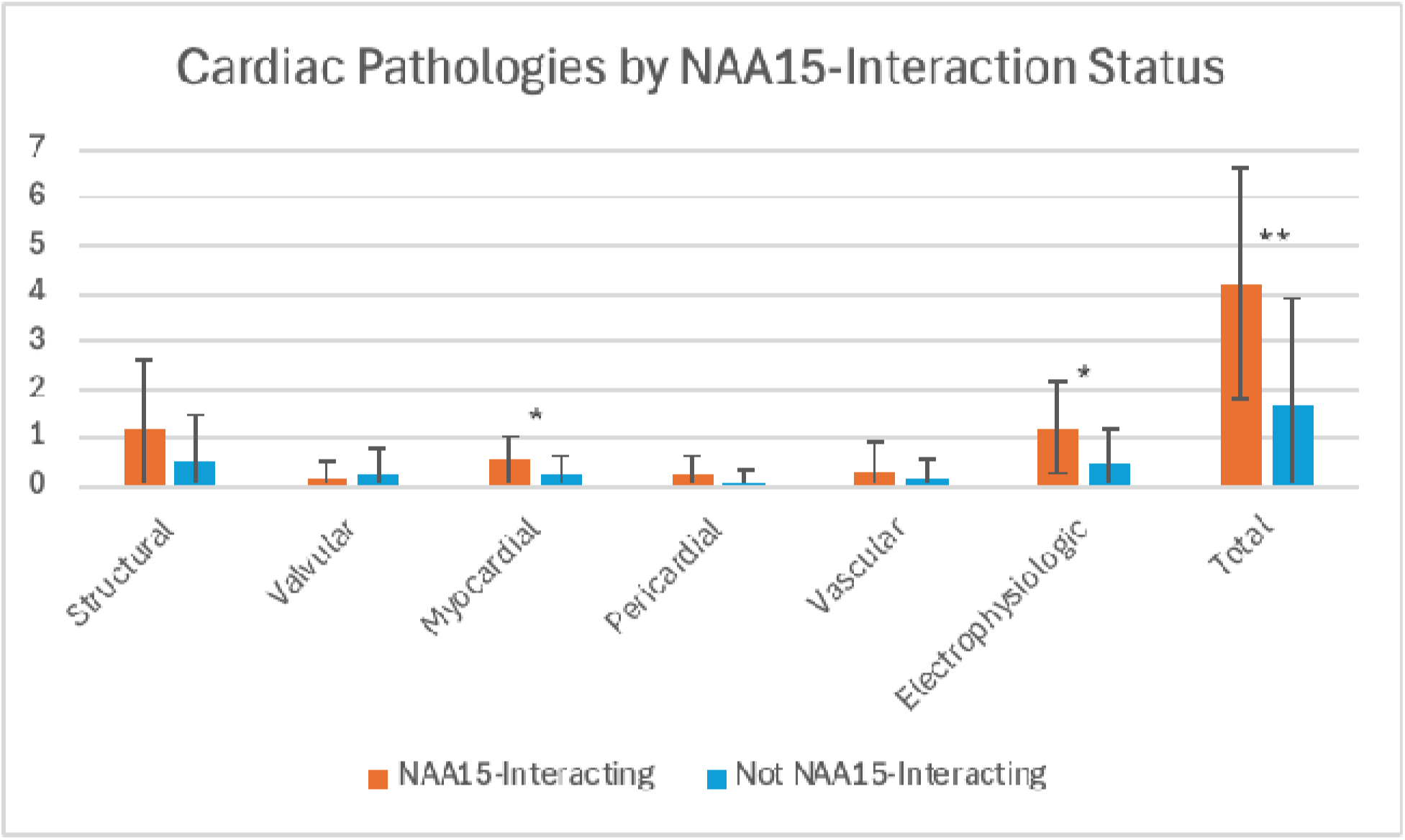
Number of Cardiac Pathologies by NAA15-Interaction Status. * p < 0.05. ** p < 0.005.

**Table 3.**
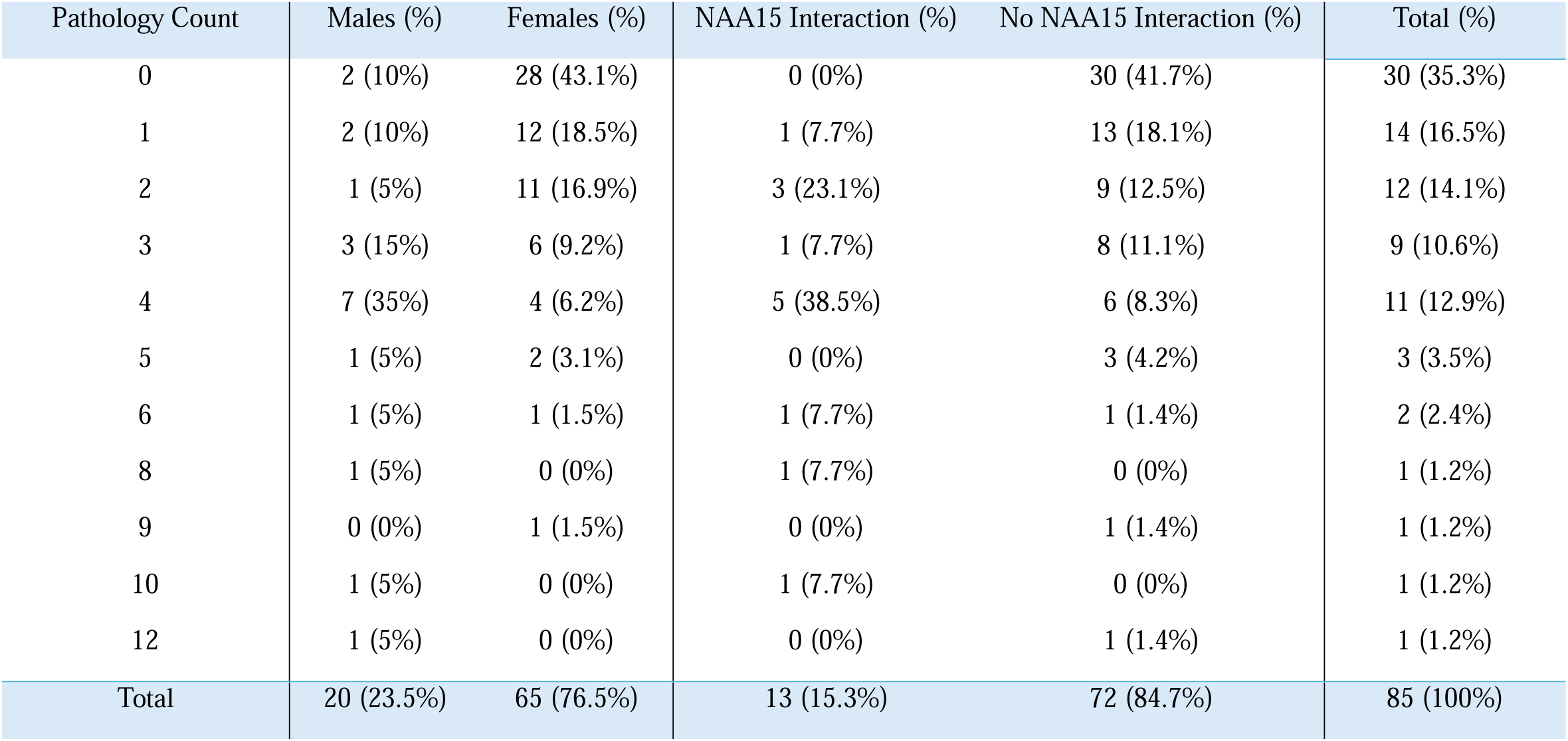
Count of Probands by Total Number of Pathology Separated by Sex and Variant. Percentages for the proportion of probands with a number of pathology are calculated using the column totals. The percentages in the total row are calculated from the total cohort (n = 85).

The average number of pathologies present per proband was 2.1 (SD = 2.4). Males, on average, had 4.1 (SD = 3.0) different cardiac abnormalities while females had 1.5 (SD = 1.8). This was statistically significant (p = .0006). Males also had a significantly greater number of myocardial abnormalities (n = 0.65; SD = 0.57) than females (n = 0.14; SD = 0.34) (p = 0.0006). In addition, males had a significantly greater number of electrophysiological abnormalities (n = 1.1; SD = 0.97) than the females (n = 0.43; SD = 0.66) (p = 0.008). Structural abnormalities were also significantly greater in the males (n = 1.1; SD = 1.3) than the females (n = 0.46; SD = 0.93) (p = .03). There was no statistically significant difference between the prevalence of specific myocardial or electrophysiologic pathologies between the sexes. There was also a significantly greater proportion of males (n = 0.35; SD = 0.48) that had cardiac arrest than the females (n = 0.015; SD = 0.12) (p = 0.003). The average number of cardiac pathologies present per group can be found in **Table 4**. This is further broken down by pathology in each group in **Supplemental Table 3**.

**Table 4.** Average Number of Cardiac Pathology by Sex.

| Mean (SD) | Structural | Valvular | Myocardial | Pericardial | Vascular | Electrophysiologic | Total |
| --- | --- | --- | --- | --- | --- | --- | --- |
| Total | 0.61 (1.1) | 0.22 (0.54) | 0.26 (0.46) | 0.11 (0.31) | 0.15 (0.47) | 0.58 (0.79) | 2.1 (2.4) |
| Male | 1.1 (1.3) | 0.15 (0.36) | 0.65 (0.57) | 0.15 (0.36) | 0.4 (0.8) | 1.1 (0.97) | 4.1 (3.0) |
| Female | 0.46 (0.93) | 0.20 (0.56) | 0.14 (0.35) | 0.09 (0.29) | 0.08 (0.27) | 0.43 (0.66) | 1.5 (1.8) |
| Female Arg83Cys (R83C) | 0.72 (1.17) | 0.31 (0.69) | 0.22 (0.42) | 0.06 (0.25) | 0.09 (0.3) | 0.66 (0.75) | 2.1 (2.1) |
| Female Other (Other) | 0.21 (0.55) | 0.18 (0.46) | 0.06 (0.24) | 0.12 (0.33) | 0.06 (0.24) | 0.21 (0.48) | 0.85 (1.2) |
| p-values | 0.03* | 0.3 | 0.0006* | 0.3 | 0.05 | 0.008* | 0.0006* |

\* Denotes significance with $\alpha < .05$ . Comparisons made between the male and female cohort.

Females were further stratified into Arg83Cys variants (R83C; n = 32) and non-R83C variants (n = 33). These groups were compared to each other and to the males to determine if the significant differences between the sexes remains when accounting for the R83C probands. The average prevalence in R83C female probands of cardiac pathology was 2.13 (SD = 2.1) versus 0.85 in non-R83C females (SD = 1.2). This difference was statistically significant (p = 0.004).

There was also a significant difference between the males and both R83C females (p = 0.016) and non-R83C females (p = 0.0002). The average prevalence of structural cardiac pathology in R83C females was 0.72 (SD = 1.17) compared to 0.21 (SD = 0.55) in non-R83C females. There was also a significant difference between the males and the non-R83C females (p = 0.01). The average prevalence in R83C female probands of myocardial pathology was 0.22 (SD = 0.42), while non-R83C females had 0.06 (SD = 0.24). There was a significant difference between the males and R83C (p = 0.01) and the non-R83C groups (p = 0.0003). The average prevalence of electrophysiologic pathology in R83C female probands was 0.66 (SD = 0.75) and 0.21 (SD = 0.48) in non-R83C females. This was a statistically significant difference (p = 0.01). There was also a significant difference between the males and the non-R83C group (p = 0.002). The remainder of the findings were not statistically significant. A summary of the mean number of cardiac pathologies present in the R83C and non-R83C groups can be found in **Table 4**. The p-values for comparisons between the groups can be found in **Table 5**.

**Table 5.** Cardiac Pathology by Males vs. Arg83Cys Females vs. Other Variant Females.

| P Values | Structural | Valvular | Myocardial | Pericardial | Vascular | Electrophysiologic | Total |
| --- | --- | --- | --- | --- | --- | --- | --- |
| M vs R83C | 0.31 | 0.28 | 0.01* | 0.35 | 0.12 | 0.14 | 0.016* |
| M vs Other | 0.01* | 0.78 | 0.0003* | 0.78 | 0.09 | 0.002* | 0.0002* |
| R83C vs Other | 0.03 | 0.38 | 0.07 | 0.42 | 0.62 | 0.01* | 0.004* |

\* Denotes significance with $\alpha < .02$

There were 13 probands with variants within the NAA15 interaction domain of NAA10 (males = 9, females = 4) and 72 without (males = 11, females = 61). There was a significant difference in the distribution of males and females who had variants within the NAA15 interaction domain (p = 0.0003). NAA15 interacting variants had an average of 4.2 (SD = 2.4) cardiac abnormalities versus 1.7 (SD = 2.2) in non-interacting variants. This difference was significant (p = 0.002). NAA15 interacting variants had 0.54 (SD = 0.50) myocardial and 1.15 (SD = 0.95) electrophysiological abnormalities versus 0.21 (SD = 0.44) myocardial and 0.47 (SD = 0.71) electrophysiological abnormalities in non-interacting variants. These differences were both statistically significant (p = 0.02). The remaining comparisons were not statistically significant. A summarization of the average number of cardiac pathologies present between NAA15 interaction and non-interacting variants and p-values can be found in **Table 6**. A breakdown and comparison of each individual cardiac pathology present in probands with and without variants in the NAA15 interaction domain can be found in **Supplemental Table 4**.

**Table 6.** Cardiac Pathology by NAA15 Interacting and Non-Interacting Variants.

| Mean (SD) | Structural | Valvular | Myocardial | Pericardial | Vascular | Electrophysiologic | Total |
| --- | --- | --- | --- | --- | --- | --- | --- |
| NAA15 Interaction (n = 13) | 1.2 (1.4) | 0.15 (0.36) | 0.54 (0.5) | 0.23 (0.42) | 0.31 (0.61) | 1.2 (0.95) | 4.2 (2.4) |
| No NAA15 Interaction (n = 72) | 0.51 (0.97) | 0.24 (0.57) | 0.21 (0.44) | 0.08 (0.28) | 0.13 (0.44) | 0.47 (0.71) | 1.7 (2.2) |
| p-value | 0.08 | 0.4 | 0.02* | 0.1 | 0.2 | 0.02* | 0.002* |

\* Denotes significance with $\alpha < .05$

Correlation analysis was also performed to determine the correlation between residue location along the primary structure of NAA10 and the presence of cardiac pathology. There was a statistically significant mild-moderate negative correlation between residue position and presence of cardiac pathology (r = -0.35; p = 0.001), a statistically significant mild negative correlation between residue position and presence of myocardial pathology (r = -0.28; p = 0.01), and a statistically significant mild-moderate negative correlation between residue position and presence of electrophysiological pathology (r = -0.36; p = 0.0008). Correlations between residue position and structural (r = -0.15), valvular (r = -0.035), pericardial (r = -0.014), and vascular (r = -0.093) pathologies were all negative, weak, and not statistically significant.

Out of 85 probands, 39 had electrophysiological data. Of the 39 with electrophysiological data, 11 had more than three reported EKGs. A total of 166 EKGs were compiled with an average age of acquisition of 22.7 (SD = 16.7) years old. The average and standard deviation of the age of EKG acquisition, rate, PR, QRS, QT, and QTc values for the cohort and probands with more than 3 recorded EKGs can be seen in **Table 7**.

**Table 7.**
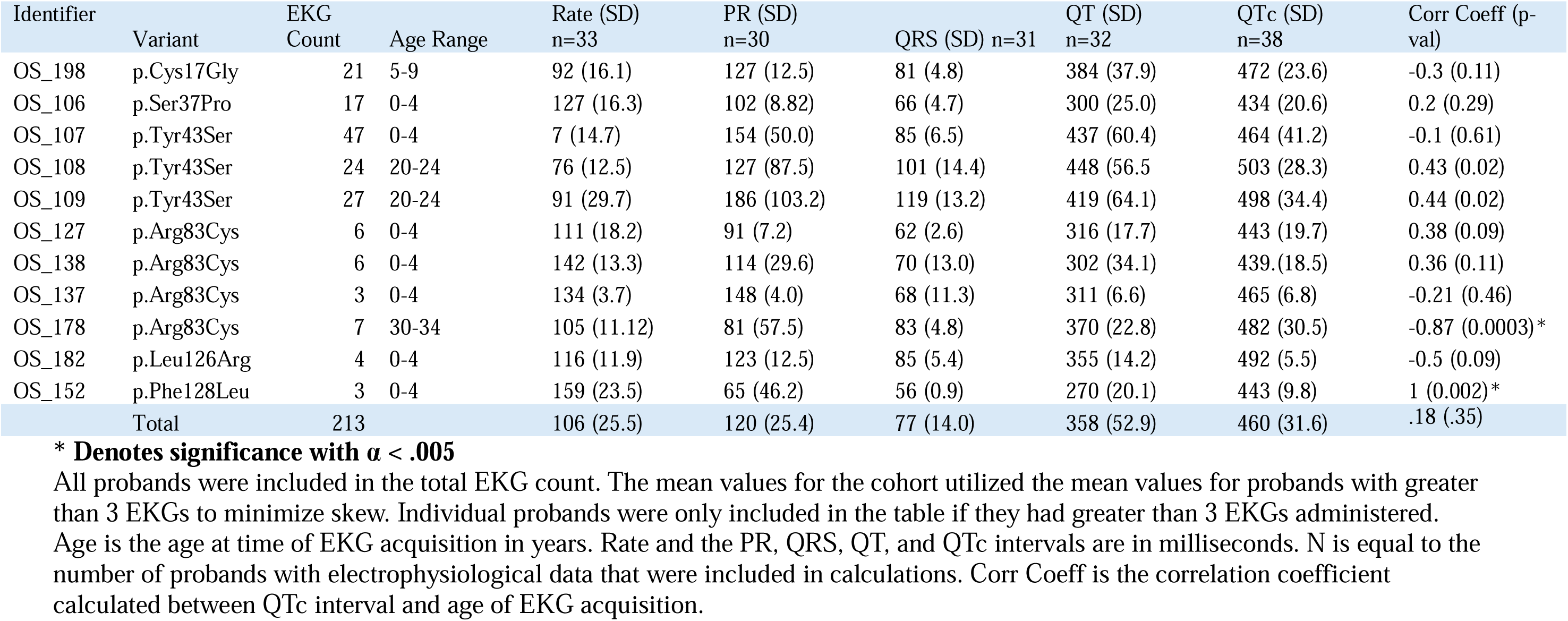
Average EKG Parameters by Cohort and Individual Proband.

| Identifier | Variant | EKG<br>Count | Age Range | Rate (SD)<br>n=33 | PR (SD)<br>n=30 | QRS (SD) n=31 | QT (SD)<br>n=32 | QTc (SD)<br>n=38 | Corr Coeff (p-<br>val) |
| --- | --- | --- | --- | --- | --- | --- | --- | --- | --- |
| OS_198 | p.Cys17Gly | 21 | 5-9 | 92 (16.1) | 127 (12.5) | 81 (4.8) | 384 (37.9) | 472 (23.6) | -0.3 (0.11) |
| OS_106 | p.Ser37Pro | 17 | 0-4 | 127 (16.3) | 102 (8.82) | 66 (4.7) | 300 (25.0) | 434 (20.6) | 0.2 (0.29) |
| OS_107 | p.Tyr43Ser | 47 | 0-4 | 7 (14.7) | 154 (50.0) | 85 (6.5) | 437 (60.4) | 464 (41.2) | -0.1 (0.61) |
| OS_108 | p.Tyr43Ser | 24 | 20-24 | 76 (12.5) | 127 (87.5) | 101 (14.4) | 448 (56.5) | 503 (28.3) | 0.43 (0.02) |
| OS_109 | p.Tyr43Ser | 27 | 20-24 | 91 (29.7) | 186 (103.2) | 119 (13.2) | 419 (64.1) | 498 (34.4) | 0.44 (0.02) |
| OS_127 | p.Arg83Cys | 6 | 0-4 | 111 (18.2) | 91 (7.2) | 62 (2.6) | 316 (17.7) | 443 (19.7) | 0.38 (0.09) |
| OS_138 | p.Arg83Cys | 6 | 0-4 | 142 (13.3) | 114 (29.6) | 70 (13.0) | 302 (34.1) | 439.(18.5) | 0.36 (0.11) |
| OS_137 | p.Arg83Cys | 3 | 0-4 | 134 (3.7) | 148 (4.0) | 68 (11.3) | 311 (6.6) | 465 (6.8) | -0.21 (0.46) |
| OS_178 | p.Arg83Cys | 7 | 30-34 | 105 (11.12) | 81 (57.5) | 83 (4.8) | 370 (22.8) | 482 (30.5) | -0.87 (0.0003)* |
| OS_182 | p.Leu126Arg | 4 | 0-4 | 116 (11.9) | 123 (12.5) | 85 (5.4) | 355 (14.2) | 492 (5.5) | -0.5 (0.09) |
| OS_152 | p.Phe128Leu | 3 | 0-4 | 159 (23.5) | 65 (46.2) | 56 (0.9) | 270 (20.1) | 443 (9.8) | 1 (0.002)* |
| Total |  | 213 |  | 106 (25.5) | 120 (25.4) | 77 (14.0) | 358 (52.9) | 460 (31.6) | .18 (.35) |

**\* Denotes significance with $\alpha < .005$**
All probands were included in the total EKG count. The mean values for the cohort utilized the mean values for probands with greater
than 3 EKGs to minimize skew. Individual probands were only included in the table if they had greater than 3 EKGs administered.
Age is the age at time of EKG acquisition in years. Rate and the PR, QRS, QT, and QTc intervals are in milliseconds. N is equal to the
number of probands with electrophysiological data that were included in calculations. Corr Coeff is the correlation coefficient
calculated between QTc interval and age of EKG acquisition.

Of the 32 probands with QT and QRS interval data, 7 had variants in the NAA15 interaction domain. The average QT interval in those variants was 395 ms (SD = 45.7 ms). The average QT interval in those with variants outside of the NAA15 interaction domain was 349 ms (SD = 47.7 ms). This difference was not statistically significant (p = 0.056). Additionally, there were no statistically significant differences in the average rates (p = 0.25), PR (p = 0.18), QRS (p = 0.14), or QTc (p = 0.48) intervals between probands with variants within the NAA15 interaction domain and those with variants outside of it. Lastly, there were no statistically significant differences between male and female mean electrophysiological data. A summary of the mean EKG parameters and their p-values broken down by sex, NAA15 interaction domain, and living status are recorded in **Supplemental Table 5**.

When assessing whether probands ever had a recorded prolonged QTc interval, 85.7% (n = 6/7) of the probands with variants within the NAA15 interaction domain had at least one. For probands with variants outside of the NAA15 domain, only 51.6% (n = 16/31) probands had a recorded prolonged QTc. This was a statistically significant difference (p = 0.049). Of the males, 88.9% (n = 8/9) had at least one prolonged QTc value compared to only 48.3% of the females. This was also statistically significant (p = 0.015).

The average QTc in this cohort was 473 ms (SD = 38.7 ms). OS_178 had a strongly negative relationship between the age of EKG acquisition and QTc interval (r = -0.87). This was a statistically significant correlation (p = 0.0003). Conversely, OS_152 had a strongly positive relationship between age of acquisition and QTc interval (r = 1.0; p = .002). These relationships over time can be seen graphically in **Figure 4a** for the cohort as a whole and **Figure 4b** for each individual proband. The remainder of the correlation coefficients for the cohort and each individual proband and their p-values are present in **Table 7**.

**Figure 4.**
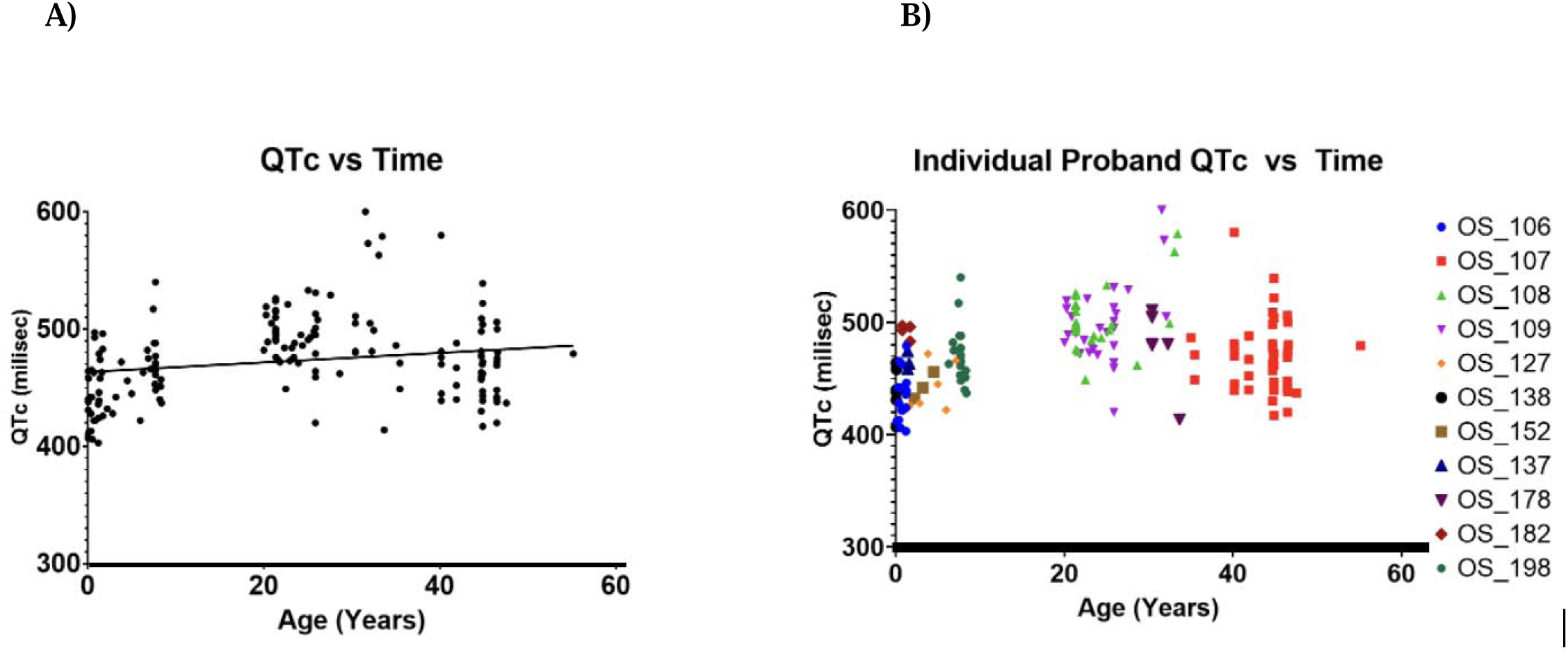
QTc Length over Time. A) QTc (milliseconds) versus age (years) for the entire cohort (n = 166 EKGs). B) QTc (milliseconds) versus age (years) for probands with greater than 3 EKGs (n = 11 probands) taken. A line of best fit was added to 2A) to aid in data visualization.

Out of 85 probands, 79 provided medication records. Of these probands, 40 were treated for noncardiac comorbidities, including seizures, behavioral disorders, intellectual disabilities, and/or infections. From the cohort, 24.05% (n = 19/79) were prescribed antiepileptics, 5.06% (n = 4/79) were on antipsychotics, 6.33% (n = 5/79) were on anxiolytics, 11.39% (n = 9/79) were on antihypertensives or stimulants, 10.13% (n = 8/79) were on antihistamines, antacids, or anti-inflammatory medication, and 12.66% (n =10/79) were on antimicrobial, antibiotic, or antifungal medication. A table of all medications and their classes can be found in **Supplemental Table 6**.

## Discussion

Cardiac issues are present across males and females with Ogden Syndrome regardless of genetic variant. Structural and electrophysiologic pathology are the most prevalent. One explanation for the structural pathology seen in OS could be due to NatA playing a role in the proper formation of the heart. NAA10 is expressed relatively consistently within the cardiac tissue of mice during development^55^ and has been shown to play a role in neuronal development^50^. Thus, dysfunction could lead to the structural malformations seen in the disease. However, human cardiac organoid studies identifying and comparing the expression of NAA10 at different stages of development have not been performed.

The incidence of electrophysiological abnormalities could be due to decreased NAA10 activity, which could contribute to altered cardiomyocyte ion channel and conductance development^42^. For example, increased calcium conductance is seen in the p.R4S, p.S37P, and p.Y43S variants of disease^46,47^. Calcium is traditionally associated with mediating excitation-coupling and cardiac contractility^51^ and increased calcium conductance can precipitate long QT syndromes^52^. In addition to increased calcium conductance, OS has been associated with increased voltage-gated sodium conductance and decreased potassium repolarizing current^42^.

Impaired repolarization is another mechanism by which long QT syndrome can develop, as seen when applying potassium blocking drugs to myocardial tissue.^53^ Due to the prevalence of prolonged QT intervals in patients with OS, it should be recommended that clinicians proceed with caution before prescribing any QT-prolonging therapies. These circumstances also warrant additional EKG monitoring and regular follow-up. Furthermore, given the lethality of ventricular fibrillation secondary to prolonged QT intervals, there should be a decreased threshold for the implantation of cardioverter defibrillation devices in this population^35^. These recommendations are extrapolated from the studies performed on cardiomyocytes with p.R4S, p.S37P, and p.Y43S variants to the rest of the cohort.

Males with OS had more severe mortality and morbidity than females, as evidenced by the greater average prevalence of myocardial, electrophysiologic, and total cardiac pathologies and greater number of cardiac arrests. This finding could be due to skewed X-inactivation in the females leading to more expression of functional NAA10 and thus a less severe phenotype^54,55^. However, the worse cardiac presentation in the males contrasts with the neurodevelopmental and ophthalmic findings of the disease, where females tended to have more severe phenotypes^29^.

While additional studies need to be performed to ascertain the pathophysiology of OS, providers should give caregivers of children with OS anticipatory guidance surrounding myocardial and arrhythmogenic disease. Additionally, it is recommended that asymptomatic female carriers of OS alleles be screened for cardiac pathology in the same way that females carrying Duchenne muscular dystrophy genes are screened to aid in early identification of cardiomyopathy^56,57^.

Interestingly, when controlling for the most prevalent variant Arg83Cys, the difference between total pathologies remained and there appeared to be a stratification of the differences between the male, Arg83Cys, and non-Arg83Cys groups. This is different than what has been observed in the neurodevelopmental symptoms of OS, in which there were no differences in adaptive functioning between probands with Arg83Cys variants and those with non-Arg83Cys variants^58^. Mechanistic studies have shown that Arg83Cys variants have reduced NAA10 activity, suggesting that reduced overall NAA10 activity may lead to different phenotypes than variants with absent NAA10 activity or those with variants within the various auxiliary binding domains of NAA10^56^.

*NAA15*-related neurodevelopmental syndrome is a disease that is related to OS in that both associated proteins are related to the NatA complex. NAA15-related neurodevelopmental syndrome has also been associated with hypertrophic obstructive cardiomyopathy (HOCM), suggesting NAA15 has a role in NatA’s function in cardiac homeostasis^44^. Characterization of cardiac ion channels of iPSC’s modeling OS variants p.S37P and p.Y43S both exhibited a long QT phenotype and the increased prevalence of electrophysiologic abnormalities in this group further supports this assertion. Our findings suggest providers should lower their threshold for referring patients with OS NAA15 interacting variants to pediatric cardiology for workup if symptomatic.

Several medications prescribed to probands in our cohort are associated with QT interval prolongation. These include levetiracetam, risperidone, lisdexamfetamine, methylphenidate, famotidine, ciprofloxacin, erythromycin, or ofloxacin ^56^. As patients with Ogden Syndrome are predisposed to cardiac issues, caution should be taken when prescribing these and similar medications. Alternative medications with lower incidence of QT interval prolongation are recommended to avoid the exacerbation of cardiac pathologies. In instances where QT-prolonging medications are necessary, there should be a low threshold for cardiac screenings, follow-up, and specialist referrals.

Lastly, due to the retrospective nature of this study, the ultra-rare nature of the disease, and the relative paucity of structural and biochemical data regarding NAA10, there are several limitations to this study. For example, while this study represents one of the largest cohorts of OS studied to date, the retrospective design leads to inconsistencies in the tests ordered between patients. This can be most clearly observed in the number of EKGs ordered, as some patients had nearly 40 times more EKGs done than others. A future study that is performed prospectively and longitudinally, ideally from birth, would help increase the internal validity of the findings. In addition, small sample size due to the ultra-rare nature of the disease acted as a limitation to our study. For example, both the male group and the NAA15 interaction group had worse cardiac outcomes than the female and non-NAA15 interaction groups. However, our cohort included a significantly greater number of males within it that have NAA15 variants. Due to the small overall number of probands, it is difficult to determine whether sex or variant type had a greater effect on cardiac outcomes in the disease. Increasing the cohort size would allow for subtle differences that were otherwise hidden to be elucidated. Lastly, the lack of biochemical and structural data regarding NAA10 and NatA function hindered our ability to provide mechanistic explanations for our clinical observations. While a handful of studies exist that helped showcase, for example, electrophysiology of cardiomyocytes in iPSC lines of several OS variants, further work must be done to characterize the NAA10 structure and perform additional electrophysiological and biochemical studies *in-vitro* to aid in understanding NAA10’s function in cardiac development.

## Conclusion

There is a high incidence of structural and electrophysiological abnormalities in OS. These issues are especially prevalent in males and in individuals with variants within the NAA15 binding domain of NAA10. Patients with OS should receive a full battery of cardiac testing upon diagnosis including echocardiography for structural abnormalities and routine monitoring of electrophysiologic parameters with EKG. Furthermore, caution should be exercised in prescribing QT-prolonging drugs to these patients. This is especially important given the high prevalence of comorbidities such as intellectual disability and seizures which often rely on therapies associated with QT prolongation. Given the ultra-rare nature of this disease, further prospective work with larger cohorts is required to more accurately determine whether sex or variant type has a greater impact on overall cardiac pathology and function.

## Supplemental Information

**S1. Additional Tables and Figures**

## Author Contributions

GJL conducted all virtual interviews with participants, with data curation conducted by EM, RP, and RO. RM, RP, and GJL were responsible for project conception. RM was responsible for data analysis. The first draft of the manuscript was written by RM and RP, with critical revision performed by GJL and RO at several points.

## Acknowledgments

We thank the families and the foundation, Ogden CARES for their participation and support. We thank Ellen Israel for assistance with early data collection for the project.

## Ethical Approval

Both oral and written patient consent were obtained for research and publication, with approval of protocol #7659 for the Jervis Clinic by the New York State Psychiatric Institute Institutional Review Board.

## Data Availability

All data are deidentified to protect subject privacy, and the underlying data cannot be shared due to these same privacy restrictions.

## References

1. Arnesen, T. et al. Proteomics analyses reveal the evolutionary conservation and divergence of N-terminal acetyltransferases from yeast and humans. Proc. Natl. Acad. Sci. U. S. A. 106, 8157–8162 (2009).

2. Van Damme, P. et al. Proteome-derived Peptide Libraries Allow Detailed Analysis of the Substrate Specificities of Nα-acetyltransferases and Point to hNaa10p as the Post-translational Actin Nα-acetyltransferase. Mol. Cell. Proteomics 10, M110.004580 (2011).

3. Aksnes, H., Ree, R. & Arnesen, T. Cotranslational, Posttranslational, and Noncatalytic Roles of N-terminal Acetyltransferases. Mol. Cell 73, 1097–1114 (2019).

4. Dörfel, M. J. & Lyon, G. J. The biological functions of Naa10 - From amino-terminal acetylation to human disease. Gene 567, 103–131 (2015).

5. Polevoda, B. & Sherman, F. N-terminal acetyltransferases and sequence requirements for N-terminal acetylation of eukaryotic proteins. J. Mol. Biol. 325, 595–622 (2003).

6. Nguyen, K. T., Mun, S.-H., Lee, C.-S. & Hwang, C.-S. Control of protein degradation by N-terminal acetylation and the N-end rule pathway. Exp. Mol. Med. 50, 1–8 (2018).

7. Lee, C.-C. et al. The Role of N-α-acetyltransferase 10 Protein in DNA Methylation and Genomic Imprinting. Mol. Cell 68, 89–103.e7 (2017).

8. Starheim, K. K., Gevaert, K. & Arnesen, T. Protein N-terminal acetyltransferases: when the start matters. Trends Biochem. Sci. 37, 152–161 (2012).

9. Gottlieb, L. & Marmorstein, R. Structure of human NatA and its regulation by the Huntingtin interacting protein HYPK. Structure 26, 925–935.e8 (2018).

10. Arnesen, T. et al. Identification and characterization of the human ARD1–NATH protein acetyltransferase complex. Biochem. J. 386, 433–443 (2005).

11. Liszczak, G. et al. Molecular Basis for Amino-Terminal Acetylation by the Heterodimeric NatA Complex. Nat. Struct. Mol. Biol. 20, 1098–1105 (2013).

12. Weyer, F. A. et al. Structural basis of HypK regulating N-terminal acetylation by the NatA complex. Nat. Commun. 8, 15726 (2017).

13. Van Damme, P. Charting the N-Terminal Acetylome: A Comprehensive Map of Human NatA Substrates. Int. J. Mol. Sci. 22, 10692 (2021).

14. Wang, T. et al. Identification and characterization of essential genes in the human genome. Science 350, 1096–1101 (2015).

15. Gao, G.-B. et al. LncRNA RGMB-AS1 inhibits HMOX1 ubiquitination and NAA10 activation to induce ferroptosis in non-small cell lung cancer. Cancer Lett. 590, 216826 (2024).

16. Lee, J. Y., Lee, S. M., Lee, W. K., Park, J. Y. & Kim, D. S. NAA10 Hypomethylation is associated with particulate matter exposure and worse prognosis for patients with non-small cell lung cancer. Animal Cells Syst. (Seoul) 27, 72–82 (2023).

17. Duong, N. X. et al. NAA10 gene expression is associated with mesenchymal transition, dedifferentiation, and progression of clear cell renal cell carcinoma. Pathol. Res. Pract. 255, 155191 (2024).

18. Zhang, Z.-Y. et al. NAA10 promotes proliferation of renal cell carcinoma by upregulating UPK1B. Eur. Rev. Med. Pharmacol. Sci. 24, 11553–11560 (2020).

19. Yao, W. et al. Exosomal circ_0026611 contributes to lymphangiogenesis by reducing PROX1 acetylation and ubiquitination in human lymphatic endothelial cells (HLECs). Cell. Mol. Biol. Lett. 28, 13 (2023).

20. Wang, D. et al. iTRAQ and two-dimensional-LC-MS/MS reveal NAA10 is a potential biomarker in esophageal squamous cell carcinoma. Proteomics Clin. Appl. 16, e2100081 (2022).

21. Fang, X. et al. ARD1 stabilizes NRF2 through direct interaction and promotes colon cancer progression. Life Sci. 313, 121217 (2023).

22. Lee, C.-F. et al. hNaa10p contributes to tumorigenesis by facilitating DNMT1-mediated tumor suppressor gene silencing. J. Clin. Invest. 120, 2920–2930 (2010).

23. Zeng, Y. et al. High expression of Naa10p associates with lymph node metastasis and predicts favorable prognosis of oral squamous cell carcinoma. Tumour Biol. 37, 6719–6728 (2016).

24. Yu, M. et al. Correlation of expression of human arrest-defective-1 (hARD1) protein with breast cancer. Cancer Invest. 27, 978–983 (2009).

25. Wang, Z. et al. Inactivation of androgen-induced regulator ARD1 inhibits androgen receptor acetylation and prostate tumorigenesis. Proc. Natl. Acad. Sci. U. S. A. 109, 3053–3058 (2012).

26. Midorikawa, Y. et al. Identification of Genes Associated with Dedifferentiation of Hepatocellular Carcinoma with Expression Profiling Analysis. Jpn. J. Cancer Res. 93, 636– 643 (2002).

27. Zeng, Y. et al. Inhibition of STAT5a by Naa10p contributes to decreased breast cancer metastasis. Carcinogenesis 35, 2244–2253 (2014).

28. Le, M.-K., Vuong, H. G., Nguyen, T. T. T. & Kondo, T. NAA10 overexpression dictates distinct epigenetic, genetic, and clinicopathological characteristics in adult gliomas. J. Neuropathol. Exp. Neurol. 82, 650–658 (2023).

29. Patel, R., et al. Ophthalmic Manifestations of NAA10-Related and NAA15-Related Neurodevelopmental Syndrome: Analysis of Cortical Visual Impairment and Refractive Errors. medRxiv 2024.02.01.24302161 (2024).

30. Sandomirsky, K., Marchi, E., Gavin, M., Amble, K. & Lyon, G. J. Phenotypic variability and gastrointestinal manifestations/interventions for growth in NAA10-related neurodevelopmental syndrome. Am. J. Med. Genet. A 191, 1293–1300 (2023).

31. Lyon, G. J. et al. Expanding the phenotypic spectrum of NAA10-related neurodevelopmental syndrome and NAA15-related neurodevelopmental syndrome. Eur. J. Hum. Genet. 31, 824–833 (2023).

32. Patel, R. et al. Neuroanatomical features of NAA10- and NAA15-related neurodevelopmental syndromes. medRxiv (2024) doi:10.1101/2024.06.24.24309433.

33. Rope, A. F. et al. Using VAAST to identify an X-linked disorder resulting in lethality in male infants due to N-terminal acetyltransferase deficiency. Am. J. Hum. Genet. 89, 28–43 (2011).

34. Shishido, A. et al. A Japanese boy with NAA10-related syndrome and hypertrophic cardiomyopathy. Hum. Genome Var. 7, 23 (2020).

35. Mizuno, Y. et al. A case of NAA10-related syndrome with prolonged QTc treated with a subcutaneous implantable cardioverter defibrillator after ventricular fibrillation. CJC Pediatr Congenit Heart Dis 1, 270–273 (2022).

36. Wei, K. & Zou, C. Clinical manifestations in a Chinese girl with heterozygous de novo NAA10 variant c. 247C > T, p. (Arg83Cys): a case report. Front. Pediatr. 11, 1198906 (2023).

37. Li, F. et al. NAA10 gene related Ogden syndrome with obstructive hypertrophic cardiomyopathy: A rare case report. Medicine (Baltimore) 103, e36034 (2024).

38. Støve, S. I. et al. A novel NAA10 variant with impaired acetyltransferase activity causes developmental delay, intellectual disability, and hypertrophic cardiomyopathy. Eur. J. Hum. Genet. 26, 1294–1305 (2018).

39. Wojciechowska, K., Zie, W., Pietrzyk, A. & Lejman, M. A four-year-old girl with pathogenic variant in the NAA10 gene and precocious puberty - case report and literature review. Ann. Agric. Environ. Med. 31, 306–310 (2024).

40. Casey, J. P. et al. NAA10 mutation causing a novel intellectual disability syndrome with Long QT due to N-terminal acetyltransferase impairment. Sci. Rep. 5, 16022 (2015).

41. Ward, T. et al. Mechanisms of Congenital Heart Disease Caused by NAA15 Haploinsufficiency. Circ. Res. 128, 1156–1169 (2021).

42. Yubero, D., Martorell, L., Nunes, T., Lyon, G. J. & Ortigoza-Escobar, J. D. Neurodevelopmental Gene-Related Dystonia: A Pediatric Case with NAA15 Variant. Mov. Disord. 37, 2320–2321 (2022).

43. Cheng, H. et al. Phenotypic and biochemical analysis of an international cohort of individuals with variants in NAA10 and NAA15. Hum. Mol. Genet. 28, 2900–2919 (2019).

44. Ritter, A. et al. Variants in NAA15 cause pediatric hypertrophic cardiomyopathy. Am. J. Med. Genet. A 185, 228–233 (2021).

45. Makwana, R., Christ, C., Marchi, E., Harpell, R. & Lyon, G. J. A Natural History of NAA15-related Neurodevelopmental Disorder Through Adolescence. medRxiv 2024.04.20.24306120 (2024).

46. Belbachir, N. et al. Studying Long QT Syndrome Caused by NAA10 Genetic Variants Using Patient-Derived Induced Pluripotent Stem Cells. Circulation 148, 1598–1601 (2023).

47. Bezzerides, V. et al. Dysregulation of N-terminal acetylation causes cardiac arrhythmia and cardiomyopathy. Res. Sq. (2024) doi:10.21203/rs.3.rs-3398860/v1.

48. Bazett, H. C. An analysis of the time relations of electrocardiograms. Ann. Noninvasive Electrocardiol. 2, 177–194 (1997).

49. Lee, M.-N., Kweon, H. Y. & Oh, G. T. N-α-acetyltransferase 10 (NAA10) in development: the role of NAA10. Exp. Mol. Med. 50, 1–11 (2018).

50. Sugiura, N., Adams, S. M. & Corriveau, R. A. An evolutionarily conserved N-terminal acetyltransferase complex associated with neuronal development. J. Biol. Chem. 278, 40113–40120 (2003).

51. Eisner, D. A., Caldwell, J. L., Kistamás, K. & Trafford, A. W. Calcium and excitation-contraction coupling in the heart. Circ. Res. 121, 181–195 (2017).

52. Zhang, Q., Chen, J., Qin, Y., Wang, J. & Zhou, L. Mutations in voltage-gated L-type calcium channel: implications in cardiac arrhythmia. Channels (Austin) 12, 201–218 (2018).

53. Morissette, P., Hreiche, R. & Turgeon, J. Drug-induced long QT syndrome and torsade de pointes. Can. J. Cardiol. 21, 857–864 (2005).

54. Bader, I. et al. Severe syndromic ID and skewed X-inactivation in a girl with NAA10 dysfunction and a novel heterozygous de novo NAA10 p.(His16Pro) variant - a case report. BMC Med. Genet. 21, 153 (2020).

55. Shvetsova, E. et al. Skewed X-inactivation is common in the general female population. Eur. J. Hum. Genet. 27, 455–465 (2019).

56. Saunier, C. et al. Expanding the Phenotype Associated with NAA10-Related N-Terminal Acetylation Deficiency. Hum. Mutat. 37, 755–764 (2016).

57. Hoogerwaard, E. M. et al. Signs and symptoms of Duchenne muscular dystrophy and Becker muscular dystrophy among carriers in The Netherlands: a cohort study. Lancet 353, 2116–2119 (1999).

58. Makwana, R., Christ, C., Marchi, E., Harpell, R. & Lyon, G. J. Longitudinal adaptive behavioral outcomes in Ogden syndrome by seizure status and therapeutic intervention. Am. J. Med. Genet. A 194, e63651 (2024).56

