## Supplementary Information for "The Cardiovascular Manifestations and Management Recommendations for Ogden Syndrome"

**Supplemental Table 1. Demographic Profile of Cohort by Country of Origin**

| Country | Count |
| --- | --- |
| USA | 41 |
| UK | 13 |
| Canada | 6 |
| Germany | 4 |
| Ireland | 3 |
| Brazil | 3 |
| Australia | 3 |
| Spain | 2 |
| France | 2 |
| China | 2 |
| Dominican Republic | 1 |
| Poland | 1 |
| Netherlands | 1 |
| Finland | 1 |
| India | 1 |
| Serbia | 1 |

**Supplemental Table 2. Discrete Counts of Individual Cardiac Pathologies Present by Sex**

**2a) Structural**

|  | Patent foramen ovale (PFO) | Atrial septal defect (ASD) | Ventricular septal defect (VSD) | Patent ductus arteriosus (PDA) | Bicuspid aortic valve | Aortic root dilatation | Ascending aorta dilation | Over-riding aorta | Tetralogy of Fallot |
| --- | --- | --- | --- | --- | --- | --- | --- | --- | --- |
| Total Count | 9 | 16 | 10 | 7 | 3 | 1 | 2 | 2 | 2 |
| Male | 4 | 7 | 5 | 5 | 1 | 0 | 0 | 0 | 0 |
| Female | 5 | 9 | 5 | 2 | 2 | 1 | 2 | 2 | 2 |

**2b) Valvular**

|  | Tricuspid regurgitation (TR) | Mitral regurgitation(MR) | Pulmonic Regurgitation | Aortic stenosis | Mitral stenosis |
| --- | --- | --- | --- | --- | --- |
| Total Count | 6 | 5 | 2 | 2 | 1 |
| Male | 2 | 1 | 0 | 0 | 0 |
| Female | 4 | 4 | 2 | 2 | 1 |

**2c) Myocardial and Pericardial**

|  | Hypertrophic cardiomyopathy (HOCM) | Ventricular hypertrophy | Cardiomegaly | Pericardial effusion | Pericarditis | Apical cyst |
| --- | --- | --- | --- | --- | --- | --- |
| Total Count | 6 | 13 | 3 | 7 | 1 | 1 |
| Male | 3 | 7 | 3 | 2 | 0 | 1 |
| Female | 3 | 6 | 0 | 5 | 1 | 0 |

**2d) Vascular**

|  | Persistent left superior vena cava (PLSVC) | Pulmonary hypertension | Pulmonary vessel stenosis | Arterial stenosis | Coronary fistula |
| --- | --- | --- | --- | --- | --- |
| Total Count | 1 | 3 | 7 | 1 | 1 |
| Male | 0 | 3 | 4 | 1 | 0 |
| Female | 1 | 0 | 3 | 0 | 1 |

**2e) Electrophysiologic**

|  | Bradycardia | Shortened PR | Prolonged QT | Supraventricular tachyarrhythmias (SVT) | AV nodal re-entrant tachycardia (AVNRT) | Atrial fibrillation | Ventricular fibrillation | Ventricular tachycardia |
| --- | --- | --- | --- | --- | --- | --- | --- | --- |
| Total Count | 5 | 2 | 28 | 4 | 1 | 4 | 2 | 3 |
| Male | 1 | 1 | 9 | 2 | 0 | 3 | 2 | 3 |
| Female | 4 | 1 | 19 | 2 | 1 | 1 | 0 | 0 |

**2f) Other**

|  | Syncopal episode | Dextrocardia | Cardiac arrest | Myocardial infarction (MI) | Heart failure |
| --- | --- | --- | --- | --- | --- |
| Total Count | 3 | 1 | 8 | 1 | 1 |
| Male | 2 | 1 | 7 | 1 | 1 |
| Female | 1 | 0 | 1 | 0 | 0 |

**Supplemental Table 3. Average Number of Individual Cardiac Pathologies Present by Sex**

Note: F = female; Avg = average; SD = standard deviation

**3a) Structural**

|  | Patent foramen ovale (PFO) | Atrial septal defect (ASD) | Ventricular septal defect (VSD) | Patent ductus arteriosus (PDA) | Bicuspid aortic valve | Aortic root dilatation | Ascending aorta dilation | Over-riding aorta | Tetralogy of Fallot |
| --- | --- | --- | --- | --- | --- | --- | --- | --- | --- |
| Total Avg | 0.11 | 0.19 | 0.12 | 0.08 | 0.04 | 0.01 | 0.02 | 0.02 | 0.02 |
| SD | 0.31 | 0.39 | 0.32 | 0.27 | 0.18 | 0.11 | 0.15 | 0.15 | 0.15 |
| Male Avg | 0.20 | 0.35 | 0.25 | 0.25 | 0.05 | 0 | 0 | 0 | 0 |
| SD | 0.40 | 0.48 | 0.43 | 0.43 | 0.22 | 0 | 0 | 0 | 0 |
| F Avg | 0.08 | 0.14 | 0.08 | 0.03 | 0.03 | 0.02 | 0.03 | 0.03 | 0.03 |
| SD | 0.27 | 0.35 | 0.27 | 0.17 | 0.17 | 0.12 | 0.17 | 0.17 | 0.17 |
| p-value | 0.11 | 0.04 | 0.06 | 0.02 | 0.36 | 0.16 | 0.08 | 0.08 | 0.08 |

**3b) Valvular**

|  | Tricuspid regurgitation (TR) | Mitral regurgitation(MR) | Pulmonic Regurgitation | Aortic stenosis | Mitral stenosis |
| --- | --- | --- | --- | --- | --- |
| Total Avg | 0.07 | 0.06 | 0.02 | 0.02 | 0.01 |
| SD | 0.26 | 0.24 | 0.15 | 0.15 | 0.11 |
| Male Avg | 0.10 | 0.05 | 0 | 0 | 0 |
| SD | 0.30 | 0.22 | 0 | 0 | 0 |
| F Avg | 0.06 | 0.06 | 0.03 | 0.03 | 0.02 |
| SD | 0.24 | 0.24 | 0.17 | 0.17 | 0.12 |
| p-value | 0.31 | 0.42 | 0.08 | 0.08 | 0.16 |

**3c) Myocardial and Pericardial**

|  | Hypertrophic cardiomyopathy (HOCM) | Ventricular hypertrophy | Cardiomegaly | Pericardial effusion | Pericarditis | Apical cyst |
| --- | --- | --- | --- | --- | --- | --- |
| Total Avg | 0.07 | 0.15 | 0.04 | 0.08 | 0.01 | 0.01 |
| SD | 0.26 | 0.36 | 0.18 | 0.27 | 0.11 | 0.11 |
| Male Avg | 0.15 | 0.35 | 0.15 | 0.10 | 0 | 0.05 |
| SD | 0.36 | 0.48 | 0.36 | 0.30 | 0 | 0.22 |
| F Avg | 0.05 | 0.09 | 0.00 | 0.08 | 0.02 | 0.00 |
| SD | 0.21 | 0.29 | 0.00 | 0.27 | 0.12 | 0.00 |
| p-value | 0.12 | 0.02 | 0.04 | 0.38 | 0.16 | 0.16 |

**3d) Vascular**

|  | Persistent left superior vena cava (PLSVC) | Pulmonary hypertension | Pulmonary vessel stenosis | Arterial stenosis | Coronary fistula |
| --- | --- | --- | --- | --- | --- |
| Total Avg | 0.01 | 0.04 | 0.08 | 0.01 | 0.01 |
| SD | 0.11 | 0.19 | 0.27 | 0.11 | 0.11 |
| Male Avg | 0.00 | 0.16 | 0.2 | 0.05 | 0 |
| SD | 0.00 | 0.36 | 0.4 | 0.22 | 0 |
| F Avg | 0.02 | 0 | 0.05 | 0 | 0.02 |
| SD | 0.12 | 0 | 0.21 | 0 | 0.12 |
| p-value | 0.16 | 0.04 | 0.06 | 0.16 | 0.16 |

**3e) Electrophysiologic**

|  | Bradycardia | Shortened PR | Prolonged QT | Supraventricular tachyarrhythmias (SVT) | AV nodal re-entrant tachycardia (AVNRT) | Atrial fibrillation | Ventricular fibrillation | Ventricular tachycardia |
| --- | --- | --- | --- | --- | --- | --- | --- | --- |
| Total Avg | 0.06 | 0.02 | 0.33 | 0.05 | 0.01 | 0.05 | 0.02 | 0.04 |
| SD | 0.24 | 0.15 | 0.47 | 0.21 | 0.11 | 0.21 | 0.15 | 0.18 |
| Male Avg | 0.05 | 0.05 | 0.45 | 0.1 | 0 | 0.15 | 0.1 | 0.15 |
| SD | 0.22 | 0.22 | 0.5 | 0.3 | 0 | 0.36 | 0.3 | 0.36 |
| F Avg | 0.06 | 0.02 | 0.29 | 0.03 | 0.02 | 0.02 | 0 | 0 |
| SD | 0.24 | 0.12 | 0.45 | 0.17 | 0.12 | 0.12 | 0 | 0 |
| p-value | 0.42 | 0.26 | 0.11 | 0.17 | 0.16 | 0.06 | 0.08 | 0.04 |

**3f) Other**

|  | Syncopal episode | Dextrocardia | Cardiac arrest | Myocardial infarction (MI) | Heart failure |
| --- | --- | --- | --- | --- | --- |
| Total Avg | 0.04 | 0.01 | 0.09 | 0.01 | 0.01 |
| SD | 0.18 | 0.11 | 0.29 | 0.11 | 0.11 |
| Male Avg | 0.1 | 0.05 | 0.35 | 0.05 | 0.05 |
| SD | 0.3 | 0.22 | 0.48 | 0.22 | 0.22 |
| F Avg | 0.02 | 0 | 0.02 | 0 | 0 |
| SD | 0.12 | 0 | 0.12 | 0 | 0 |
| p-value | 0.12 | 0.16 | 0 | 0.16 | 0.16 |

**Supplemental Table 4. Average Number of Individual Cardiac Pathologies Present by Variant**

Note: NAA15 refers to NAA10 variants within the NAA15 interaction domain

**4a) Structural**

|  | Patent foramen ovale (PFO) | Atrial septal defect (ASD) | Ventricular septal defect (VSD) | Patent ductus arteriosus (PDA) | Bicuspid aortic valve | Aortic root dilatation | Ascending aorta dilation | Over-riding aorta | Tetralogy of Fallot |
| --- | --- | --- | --- | --- | --- | --- | --- | --- | --- |
| Naa15 | 0.31 | 0.23 | 0.23 | 0.31 | 0.08 | 0 | 0 | 0 | 0 |
| SD | 0.46 | 0.42 | 0.42 | 0.46 | 0.27 | 0 | 0 | 0 | 0 |
| No Naa15 | 0.07 | 0.18 | 0.10 | 0.04 | 0.03 | 0.01 | 0.03 | 0.03 | 0.03 |
| SD | 0.25 | 0.38 | 0.30 | 0.20 | 0.16 | 0.12 | 0.16 | 0.16 | 0.16 |
| p value | 0.05 | 0.35 | 0.15 | 0.04 | 0.27 | 0.16 | 0.08 | 0.08 | 0.08 |

**4b) Valvular**

|  | Tricuspid regurgitation (TR) | Mitral regurgitation(MR) | Pulmonic Regurgitation | Aortic stenosis | Mitral stenosis |
| --- | --- | --- | --- | --- | --- |
| Naa15 | 0.08 | 0.08 | 0 | 0 | 0 |
| SD | 0.27 | 0.27 | 0 | 0 | 0 |
| No Naa15 | 0.07 | 0.06 | 0.03 | 0.03 | 0.01 |
| SD | 0.25 | 0.23 | 0.16 | 0.16 | 0.12 |
| p value | 0.46 | 0.40 | 0.08 | 0.08 | 0.16 |

**4c) Myocardial and Pericardial**

|  | Hypertrophic cardiomyopathy (HOCM) | Ventricular hypertrophy | Cardiomegaly | Pericardial effusion | Pericarditis | Apical cyst |
| --- | --- | --- | --- | --- | --- | --- |
| Naa15 | 0 | 0.38 | 0.15 | 0.23 | 0 | 0 |
| SD | 0 | 0.49 | 0.36 | 0.42 | 0 | 0 |
| No Naa15 | 0.08 | 0.11 | 0.01 | 0.06 | 0.01 | 0.01 |
| SD | 0.28 | 0.31 | 0.12 | 0.23 | 0.12 | 0.12 |
| p value | 0.01 | 0.04 | 0.10 | 0.09 | 0.16 | 0.16 |

**4d) Vascular**

|  | Persistent left superior vena cava (PLSVC) | Pulmonary hypertension | Pulmonary vessel stenosis | Arterial stenosis | Coronary fistula |
| --- | --- | --- | --- | --- | --- |
| Naa15 | 0 | 0.08 | 0.23 | 0 | 0 |
| SD | 0 | 0.28 | 0.42 | 0 | 0 |
| No Naa15 | 0.01 | 0.03 | 0.06 | 0.01 | 0.01 |
| SD | 0.12 | 0.16 | 0.23 | 0.12 | 0.12 |
| p value | 0.16 | 0.26 | 0.09 | 0.16 | 0.16 |

**4e) Electrophysiologic**

|  | Bradycardia | Shortened PR | Prolonged QT | Supraventricular tachyarrhythmias (SVT) | AV nodal re-entrant tachycardia (AVNRT) | Atrial fibrillation | Ventricular fibrillation | Ventricular tachycardia |
| --- | --- | --- | --- | --- | --- | --- | --- | --- |
| Naa15 | 0.08 | 0 | 0.54 | 0 | 0 | 0.31 | 0.15 | 0.08 |
| SD | 0.27 | 0 | 0.50 | 0 | 0 | 0.46 | 0.36 | 0.27 |
| No Naa15 | 0.06 | 0.03 | 0.29 | 0.06 | 0.01 | 0 | 0 | 0.03 |
| SD | 0.23 | 0.16 | 0.45 | 0.23 | 0.12 | 0 | 0 | 0.16 |
| p value | 0.40 | 0.08 | 0.06 | 0.02 | 0.16 | 0.02 | 0.08 | 0.27 |

**4f) Other**

|  | Syncopal episode | Dextrocardia | Cardiac arrest | Myocardial infarction | Heart failure |
| --- | --- | --- | --- | --- | --- |
| Naa15 | 0.08 | 0.08 | 0.38 | 0.08 | 0 |
| SD | 0.27 | 0.27 | 0.49 | 0.27 | 0 |
| No Naa15 | 0.03 | 0 | 0.04 | 0 | 0.01 |
| SD | 0.16 | 0 | 0.20 | 0 | 0.12 |
| p value | 0.27 | 0.17 | 0.02 | 0.17 | 0.16 |

**Supplemental Table 5. Average EKG Parameters by Sex, Variant, and Mortality**

Note: Mean (1) refers to the mean of values that are positive for that variable. For example, the Mean (1) row of the NAA15 column refers to the mean value of probands with variants within the NAA15 interaction domain of NAA10. Mean (0) would be the opposite of that value.

|  | Rate | | | PR | | | QRS | | |
| --- | --- | --- | --- | --- | --- | --- | --- | --- | --- |
|  | NAA15 | Female | Deceased | NAA15 | Female | Deceased | NAA15 | Female | Deceased |
| Mean (1) | 95.2 | 103.1 | 118.7 | 133.5 | 118.8 | 112.1 | 86.3 | 75.1 | 72.6 |
| SD | 24.7 | 23.8 | 19.2 | 26.9 | 24.0 | 11.3 | 17.1 | 11.4 | 6.4 |
| Mean (0) | 108.8 | 116.2 | 104.6 | 115.9 | 124.1 | 120.9 | 74.1 | 82.6 | 77.3 |
| SD | 24.4 | 27.0 | 25.2 | 22.8 | 27.4 | 25.9 | 11.2 | 18.7 | 14.2 |
| p val | 0.26 | 0.31 | 0.41 | 0.18 | 0.68 | 0.4 | 0.14 | 0.38 | 0.43 |

|  | QT | | | QTc | | |
| --- | --- | --- | --- | --- | --- | --- |
|  | NAA15 | Female | Deceased | NAA15 | Female | Deceased |
| Mean (1) | 384.3 | 355.8 | 332.6 | 467.5 | 455.0 | 467.8 |
| SD | 54.3 | 50.1 | 37.3 | 26.2 | 28.1 | 21.5 |
| Mean (0) | 348.9 | 359.4 | 359.1 | 458.6 | 477.1 | 459.4 |
| SD | 47.8 | 55.7 | 52.0 | 32.0 | 34.5 | 32.0 |
| p val | 0.18 | 0.89 | 0.43 | 0.48 | 0.12 | 0.57 |

**Supplemental Table 6. Breakdown of Medications Prescribed to Treat Our Cohort**

| Antiepileptics | Antipsychotics | Anxiolytics | Antihypertensives or Stimulants | Antihistamines, Antacids, or Anti-inflammatory medications | Antimicrobial, Antibiotic, or Antifungal medications |
| --- | --- | --- | --- | --- | --- |
| Carbamazepine | Aripiprazole | Buspirone | Clonidine ER oral suspension | Cetirizine | Benzylpenicillin |
| Clobazam | Risperidone | Diazepam | Lisdexamfetamine | Cyproheptadine | Cefalexin/  Cephalexin |
| Epidiolex (Cannabidiol) |  | Gabapentin | Methylphenidate | Famotidine | Ceftriaxone |
| Gabapentin |  | Lorazepam | Guanfacine | Montelukast | Ciprofloxacin |
| Lacosamide |  | Pregabalin |  | Ranitidine | Clindamycin |
| Lamotrigine |  |  |  |  | Erythromycin |
| Lorazepam |  |  |  |  | Gentamicin |
| Levetiracetam |  |  |  |  | Ofloxacin |
| Oxcarbazepine |  |  |  |  | Nystatin |
| Phenobarbital |  |  |  |  |  |
| Pregabalin |  |  |  |  |  |
| Topiramate |  |  |  |  |  |
| Valproate |  |  |  |  |  |
| Vigabatrin |  |  |  |  |  |
| Zonisamide |  |  |  |  |  |
